# Public Attitudes to Implementing Financial Incentives in Stop Smoking Services in Ireland

**DOI:** 10.1101/2023.01.13.23284530

**Authors:** Ellen Cosgrave, Aishling Sheridan, Edward Murphy, Martina Blake, Rikke Siersbaek, Sarah Parker, Sara Burke, Frank Doyle, Paul Kavanagh

## Abstract

**INTRODUCTION:** Financial incentives improve stop smoking service outcomes. Views on acceptability can influence implementation success. To inform implementation planning in Ireland, public attitudes to financial incentives in stop smoking services were measured.

**METHODS:** A cross-sectional telephone survey was administered to a random digit dialled sample of 1000 people in Ireland aged 15 years and older in 2022. The questionnaire included items on support for financial incentives under different conditions. Prevalence of support was calculated with 95% Confidence Intervals (CIs) and multiple logistic regression identified associated factors using Adjusted Odds Ratios (aORs, with 95% CIs).

**RESULTS:** Almost half (47.0%, 95% CI 43.9%-50.1%) supported at least one type of financial incentive to stop smoking, with support more prevalent for shopping vouchers (43.3%, 95% CI 40.3%-46.5%) than cash payments (32.1%, 95% CI 29.2%-35.0%).

Support was similar for universal and income-restricted schemes. Of those who supported financial incentives, the majority (60.6%) believed the maximum amount given on proof of stopping smoking should be under €250 (median=€100, range=€1-€7000). Versus comparative counterparts, those of lower educational attainment (aOR 1.49 95% CI 1.10-2.03, p=0.010) and tobacco/e-cigarette users (aOR 1.43 95% CI 1.02-2.03, p=0.041) were significantly more likely to support either financial incentive type, as were younger people.

**CONCLUSIONS:** While views on financial incentives to stop smoking in Ireland were mixed, the intervention is more acceptable in groups experiencing the heaviest burden of smoking-related harm and most capacity to benefit. Engagement and communication must be integral to planning for successful implementation to improve stop smoking service outcomes.

## INTRODUCTION

Smoking continues to cause harm on a huge scale and helping people stop remains a key public health priority[1]. The components of effective stop smoking support are well-established[2,3], meaning that the challenge is effective implementation, especially for lower-income groups where the burden of smoking-related harm is greatest and for whom tailored stop smoking services have potential pro-equity impact[4]. There is high-certainty evidence that adding financial incentives to stop smoking services can improve outcomes[5], however, knowledge to guide effective implementation design is lacking[6].

A focus on ensuring success across implementation outcomes can help translate well-established research evidence on financial incentives into better stop smoking services[7]. Acceptability, which has been defined as “*the extent to which people delivering or receiving a healthcare intervention consider it to be appropriate, based on anticipated or experienced cognitive and emotional responses to the intervention across implementation stakeholders*”[8] can positively influence scalable and sustainable implementation of healthcare interventions[9]. Assessing acceptability of financial incentives for health-behaviour change is especially important since these complex interventions can evoke mixed reactions[10].

While Ireland has made good progress in reducing smoking prevalence, it faces challenges with widening social inequalities in smoking[11]. Recently published National Stop Smoking Guidelines identified financial incentives as a promising intervention to improve stop smoking services [12], especially for people in lower-income groups, but recommended further local research for effective implementation design and planning.

To inform potential implementation in stop smoking services, this study aimed to measure perceived acceptability of financial incentives among the Irish public.

## METHODS

This cross-sectional study used telephone delivery of a survey instrument to a representative sample of 1000 members of the Irish public aged 15 years and older recruited via random digit-dialling in 2022. Participants were excluded if they did not have a telephone, were non-fluent in the English language or if they did not respond to the survey completely.

A literature-informed instrument measured agreement with statements on financial incentives to stop smoking in different forms and settings. Responses were grouped as “support” (“strongly agree”/”somewhat agree”), “indifferent” (“neither agree nor disagree”/”don’t know”) and “oppose” (“somewhat disagree”/”strongly disagree”). Respondents also identified a maximum acceptable incentive value. Tobacco or e-cigarette use status and socio-demographic characteristics were also collected. The questions were embedded in a wider survey of public attitudes to tobacco endgame[13].

Prevalence of key measures were calculated with 95% Confidence Intervals (CIs), which were used to compare responses together with Chi-Square testing. Multiple logistic regression identified factors independently associated with support using Adjusted Odds Ratios (aORs) with 95% CIs. Re-weighting in line with recent population estimates for gender, age, region and social class was employed prior to all analyses. Analyses were conducted in IBM SPSS Statistics for Windows Version 26.0.

## RESULTS

Response rate was 30% (N=1,000). Almost half (47.0%, 95% CI 43.9%-50.1%) supported at least one type of financial incentive for smoking cessation, either shopping vouchers or cash payments. Support for shopping vouchers was higher than for cash payments (43.3% (95% CI 40.3%-46.5%) versus 32.1% (95% CI 29.2%-35.0%), Chi-Square Statistic 27.16, p-value < 0.00001). Approximately one-in-ten were indifferent to cash incentives (9.8%, 95% CI 8.0-11.6) and to voucher incentives (10.4%, 95% CI 8.5-12.3) respectively (Supplementary Material).

Regarding conditions, a similar proportion of respondents supported financial incentives for anyone who can prove that they have stopped smoking regardless of their income (unrestricted or universal financial incentives) as supported these only for people on low incomes (restricted financial incentives or targeting by social group) (33.0% (95% CI 29.1%- 37.0%) versus 32.1% (95% CI 28.2%-36.1%), Chi-Square Statistic 0.012, p-value=0.93).

Of those who supported financial incentives, the majority (60.5%, 95% CI 55.4%-65.4%), identified a maximum acceptable value under €250 (median=€100, range=€1-€7000).

Respondent age, gender, region of residence, social class, educational level, and tobacco/e-cigarette use status were included in the final multiple logistic regression model to identify factors independently associated with support for either incentive type. Versus comparative counterparts, those of lower educational attainment (aOR 1.49, 95% CI 1.10-2.03) and tobacco/e-cigarette users (aOR 1.43, 95% CI 1.02-2.03) were significantly more likely to support either type of financial incentive (Table 1). Respondents in older age groups were less likely to support either incentive type than younger counterparts, however, there was no association between financial incentive support and gender.

**TABLE 1:** Multiple Logistic Regression Analysis of Factors Associated with Participant Support for Financial Incentives (either cash or shopping voucher incentives)

| Characteristic | N (%) | Unadjusted OR<br>(95% CI) | Adjusted OR<br>(95% CI) | P-value |
| --- | --- | --- | --- | --- |
| **Gender** |  |  |  |  |
| Female | 532 (53.2) | 1 | 1 |  |
| Male | 468 (46.8) | 1.16 (0.91-1.49) | 1.07 (0.82-1.39) | 0.623 |
| **Age (years)** |  |  |  |  |
| 15-24 | 153 (15.3) | **1.76 (1.11-2.79)** | 1.31 (0.80-2.15) | 0.289 |
| 25-34 | 149 (14.9) | 1 | 1 |  |
| 35-44 | 181 (18.1) | **0.60 (0.39-0.92)** | **0.61 (0.39-0.94)** | **0.026** |
| 45-54 | 175 (17.5) | **0.45 (0.29-0.70)** | **0.44 (0.28-0.70)** | **<0.001** |
| 55-64 | 151 (15.1) | **0.49 (0.31-0.78)** | **0.43 (0.26-0.70)** | **0.001** |
| ≥65 | 191 (19.1) | 0.68 (0.44-1.04) | **0.57 (0.36-0.92)** | **0.020** |
| **Region** |  |  |  |  |
| Leinster | 562 (56.2) | 1 | 1 |  |
| Munster | 299 (29.9) | 0.78 (0.58-1.05) | 0.82 (0.61-1.12) | 0.214 |
| Connaught/Ulster | 139 (13.9) | 1.28 (0.91-1.80) | 1.31 (0.92-1.88) | 0.140 |
| **Social class** |  |  |  |  |
| Higher (A,B,C1) | 345 (34.5) | 1 | 1 |  |
| Lower (C2,D,E) | 619 (61.9) | 1.43 (0.83-2.46) | 1.21 (0.68-2.13) | 0.516 |
| Farmer | 36 (3.6) | 0.98 (0.57-1.69) | 1.00 (0.56-1.79) | 0.995 |
| **Educational attainment** |  |  |  |  |
| Higher | 594 (59.4) | 1 | 1 |  |
| Lower | 406 (40.6) | **1.85 (1.44-2.38)** | **1.49 (1.10-2.03)** | **0.010** |
| **Tobacco/e-cigarette user** |  |  |  |  |
| No | 827 (82.1) | 1 | 1 |  |
| Yes | 168 (16.9) | **1.67 (1.22-2.30)** | **1.43 (1.02-2.03)** | **0.041** |

aORs have been adjusted for all other characteristics in the table. CI= 95% confidence interval;
Bold=p value < 0.05; Naglekerke $r^2$ = 0.095

## DISCUSSION

Financial incentives are a relatively new innovation to improve stop smoking service effectiveness[5], and our study assessed public attitudes to this novel for the first time in Ireland. Despite potential effectiveness, we found views on the acceptability of implementing financial incentives in stop smoking services were mixed. However, the intervention was more acceptable in groups experiencing the greatest burden of smoking-related harm who have most capacity to benefit. Incentive type matters, with higher support for shopping vouchers than cash payments, although support for targeting of the financial incentive to people to low income and a universal approach was similar. Potential scale of financial incentives that would be supported has been delineated in Ireland, with values of less than €250 being most popular.

A recently updated systematic review found that public views on acceptability of financial incentives for health-related behaviour change can be polarised[10]. Our findings that vouchers were more acceptable than cash and that lower maximum incentives values are preferred are consistent with studies on acceptability of financial incentives for health-related behaviour change generally[10].

Concerns regarding fairness are a common theme in studies on public views of financial incentives acceptability[10]. In Ireland, as in many high income countries, the social patterning of smoking is increasing and leading to widening of social inequalities in health[11]. Using financial incentives to target stop smoking services improvements for lower-income groups has potential pro-equity impact[4], and is a critical implementation design decision point. In this study, support for universal financial incentives and for targeting to people with lower incomes was similar; in other studies, universal approaches were often more acceptable to the general public[10]. However, we also found that groups in Ireland with greatest need and most capacity to benefit from implementation of financial incentives in stop smoking services (younger people with lower educational attainment who smoke) were more likely to find the intervention acceptable. In other studies, acceptability was not always higher among groups with more capacity to benefit from financial incentives to help change unhealthy behaviours[14,15]. Compared to universal approaches, pursuing equity through targeting financial incentives to people with lower incomes may lead to friction or trade-offs in acceptability across stakeholders groups[10]. While this approach may evoke mixed reactions across the public generally, many of whom may not need the service, targeting financial incentives to those with lower incomes who smoke may be more acceptable in this group who urgently need improved stop smoking services to address widening health inequalities.

Tensions with fairness underline the importance of messaging, another common theme in research on acceptability of financial incentives[10]. This is the first discussion of using financial incentives to improve stop smoking services with the public in Ireland, and respondents in our study were not provided with information on general intervention rationale or specific arguments for targeting financial incentives to lower-income groups. These messages about implementation of financial incentives matter. For example, a discrete choice experiment found financial incentives acceptability increased when respondents were provided with information on increasing magnitude of effectiveness[16]. Sekhon et al identify intervention coherence and perceived effectiveness as component constructs of their theoretical framework on acceptability[8], which can usefully guide stakeholder communication and engagement for successful implementation of this novel and potentially polarising healthcare intervention. International evidence is useful, but local research is needed to inform context-specific approaches to stakeholder communication and engagement, since social context influences views on financial incentives acceptability[17], and media representation also shapes opinions of the intervention[18].

This is the first study in Ireland to measure acceptability of financial incentives in stop smoking services. Given the need to improve stop smoking services in Ireland, especially for people in lower groups experiencing widening smoking-related health inequalities, the study exemplifies the role of contextually-relevant evidence in improving planning for implementation success. It is, however, limited by the response rate and scope. It will benefit from complementary qualitative studies to provide a richer evidence on this complex challenge, which are planned.

## CONCLUSIONS

Adding financial incentives to stop smoking services can improve effectiveness. Translating current research evidence into better outcomes for those with greatest need is a complex challenge requiring careful design and planning to negotiate acceptability for implementation success. While views of the Irish public on acceptability of financial incentives to stop smoking were mixed, there was greater acceptability among groups who will benefit most from the improvement in stop smoking service effectiveness. Ongoing communication and engagement across stakeholder groups is essential for effective implementation of financial incentives in stop smoking services. Explaining rationale for potentially divisive design decisions regarding targeting to address health inequalities together with demonstration and feedback of real-world effectiveness are important considerations. Careful piloting involving implementation stakeholders is planned in Ireland prior to scaling and provides an opportunity to build more widespread support to sustain successful implementation of financial incentives for better stop smoking services..

## Data Availability

All data produced in the present study are available upon reasonable request to the authors

## DECLARATION OF INTERESTS

The authors have each completed and submitted an ICMJE Form for Disclosure of Potential Conflicts of Interest. The authors declare that they have no competing interests, financial or otherwise, related to the current work.

## FUNDING

Research reported was funded by the HSE Tobacco Free Ireland Programme.

## ACKNOWLEDGEMENTS

None

## AUTHORS’ CONTRIBUTIONS

EC and PK made substantial contributions to the conception and design of the work; and to the acquisition, analysis, or interpretation of data for the work.

AS, EM and MB made substantial contributions to the conception and design of the work and the acquisition of data for the work.

RS, SP, SB and FD made substantial contributions to the conception and design of the work. All authors finally approval of the version to be published.

All authors agree to be accountable for all aspects of the work in ensuring that questions related to the accuracy or integrity of any part of the work are appropriately investigated and resolved.

## SUPPLEMENTARY MATERIALS

**Table:** Support for Financial Incentives According to Tobacco/E-cigarette Use Status (N=1,000)

| Measure | Total<br>n (%) | *Tobacco/e-*<br>*cigarette User*<br>n (%) | *Non-Tobacco/e-*<br>*cigarette User*<br>n (%) |
| --- | --- | --- | --- |
| **“Shopping vouchers should be provided to people who prove that they have stopped smoking”** |  |  |  |
| Agree | 434 (43.4) | 96 (49.7) | 337 (42.0) |
| Indifferent | 98 (9.8) | 20 (10.4) | 78 (9.7) |
| Disagree | 469 (46.9) | 77 (39.9) | 388 (48.3) |
| **“A cash payment should be provided to people who prove that they have stopped smoking”** |  |  |  |
| Agree | 321 (32.1) | 79 (41.1) | 242 (30.2) |
| Indifferent | 104 (10.4) | 22 (11.4) | 82 (10.2) |
| Disagree | 575 (57.5) | 92 (47.7) | 477 (59.6) |

## Notes

### Competing Interest Statement

The authors have declared no competing interest.

### Author Declarations

Royal College of Physicians in Ireland (RCPI) Ethics Committee (RECAP 157)

## REFERENCES

1. World Health Organization. WHO report on the global tobacco epidemic, 2019: offer help to quit tobacco use. https://apps.who.int/iris/handle/10665/325968 Published 2019. Accessed January 4, 2023.

2. Hartmann-Boyce J, Livingstone-Banks J, Ordóñez-Mena JM, et al. Behavioural interventions for smoking cessation: an overview and network meta-analysis. Cochrane Database Syst Rev. 2021;1:CD013229. Published 2021 Jan 4. doi:10.1002/14651858.CD013229.pub2

3. Hartmann-Boyce J, Livingstone-Banks J, Ordóñez-Mena JM, et al. Behavioural interventions for smoking cessation: an overview and network meta-analysis. Cochrane Database Syst Rev. 2021;1:CD013229. Published 2021 Jan 4. doi:10.1002/14651858.CD013229.pub2

4. Smith CE, Hill SE, Amos A. Impact of population tobacco control interventions on socioeconomic inequalities in smoking: a systematic review and appraisal of future research directions [published online ahead of print, 2020 Sep 29]. Tob Control. 2020;30(e2):e87–e95. doi:10.1136/tobaccocontrol-2020-055874

5. Notley C, Gentry S, Livingstone-Banks J, Bauld L, Perera R, Hartmann-Boyce J. Incentives for smoking cessation. Cochrane Database Syst Rev. 2019;7(7):CD004307. Published 2019 Jul 17. doi:10.1002/14651858.CD004307.pub6

6. Miranda JJ, Pesantes MA, Lazo-Porras M, et al. Design of financial incentive interventions to improve lifestyle behaviors and health outcomes: A systematic review. Wellcome Open Res. 2021;6:163. Published 2021 Sep 2. doi:10.12688/wellcomeopenres.16947.2

7. Proctor E, Silmere H, Raghavan R, et al. Outcomes for implementation research: conceptual distinctions, measurement challenges, and research agenda. Adm Policy Ment Health. 2011;38(38):65–76. doi:10.1007/s10488-010-0319-7

8. Sekhon M, Cartwright M, Francis JJ. Acceptability of healthcare interventions: an overview of reviews and development of a theoretical framework. BMC Health Serv Res. 2017;17(17):88. Published 2017 Jan 26. doi:10.1186/s12913-017-2031-8

9. Klaic M, Kapp S, Hudson P, et al. Implementability of healthcare interventions: an overview of reviews and development of a conceptual framework. Implement Sci. 2022;17(17):10. Published 2022 Jan 27. doi:10.1186/s13012-021-01171-7

10. Hoskins K, Ulrich CM, Shinnick J, Buttenheim AM. Acceptability of financial incentives for health-related behavior change: An updated systematic review. Prev Med. 2019;126:105762. doi:10.1016/j.ypmed.2019.105762

11. Health Service Executive Tobacco Free Ireland Programme. State of Tobacco Control in Ireland, 2022. https://www.hse.ie/eng/about/who/tobaccocontrol/news/state-of-tobacco-control-report-2022.pdf. Published May, 2022. Accessed January 4, 2023.

12. National Clinical Effectiveness Committee, Department of Health in Ireland. Stop Smoking, National Clinical Guideline 28. https://www.gov.ie/en/publication/4828b-stop-smoking/ Published January, 2022. Accessed January 4, 2023.

13. Is the Public Ready for a Tobacco-Free Ireland? A National Survey of Public Knowledge and Attitudes to Tobacco Endgame in Ireland. Ellen Cosgrave, Martina Blake, Edward Murphy, Aishling Sheridan, Frank Doyle, Paul Kavanagh. medRxiv 2022.12.01.22282993; doi: https://doi.org/10.1101/2022.12.01.22282993

14. Giles, E. L., Becker, F., Ternent, L., Sniehotta, F. F., McColl, E., & Adams, J. (2016). Acceptability of Financial Incentives for Health Behaviours: A Discrete Choice Experiment. PloS one, 11(6), e0157403. https://doi.org/10.1371/journal.pone.0157403

15. Hoddinott P, Morgan H, MacLennan G, et al. Public acceptability of financial incentives for smoking cessation in pregnancy and breast feeding: a survey of the British public. BMJ Open. 2014;4(4):e005524. Published 2014 Jul 18. doi:10.1136/bmjopen-2014-005524

16. Promberger M, Dolan P, Marteau TM. “Pay them if it works”: discrete choice experiments on the acceptability of financial incentives to change health related behaviour. Soc Sci Med. 2012;75(75):2509–2514. doi:10.1016/j.socscimed.2012.09.033

17. Berlin N, Goldzahl L, Bauld L, Hoddinott P, Berlin I. Public acceptability of financial incentives to reward pregnant smokers who quit smoking: a United Kingdom-France comparison. Eur J Health Econ. 2018;19(19):697–708. doi:10.1007/s10198-017-0914-6

18. Parke H, Ashcroft R, Brown R, Marteau TM, Seale C. Financial incentives to encourage healthy behaviour: an analysis of U.K. media coverage. Health Expect. 2013;16(16):292–304. doi:10.1111/j.1369-7625.2011.00719.x

